# Recent update on COVID-19 in India: Is locking down the country enough?

**DOI:** 10.1101/2020.04.06.20053124

**Authors:** Jitender Singh Virk, Syed Azmal Ali, Gurjeet Kaur

## Abstract

**Background:** India is the second-largest population in the world, and it is not well equipped, hitherto, in the scenario of the global pandemic, SARS-CoV-2 could impart a devastating impact on the Indian population. Only way to respond against this critical condition is by practicing large-scale social distancing. India lock down for 21 days, however, till 7 April 2020, SARS-CoV-2 positive cases were growing exponentially, which raises the concerns if the number of reported and actual cases are similar.

**Methods:** We use Lasso Regression with *α* = 0.12 and Polynomial features of degree 2 to predict the growth factor. Also, we predicted Logistic curve using the Prophet Python. Further, using the growth rate to logistic, and carrying capacity is 20000 allowed us to calculate the maximum cases and new cases per day.

**Results:** We found the predicted growth factor with a standard deviation of 0.3443 for the upcoming days. When the growth factor becomes 1.0, which is known as Inflection point, it will be safe to state that the rate is no longer exponential. The estimated time to reach the inflection point is between 15-20 April. At that time, the estimated number of total positive cases will be over 12500, if lockdown remains continue.

**Conclusions:** Our analysis suggests that there is an urgent need to take action to extend the period of lockdown and allocate enough resources, including personnel, beds, and intensive care facilities, to manage the situation in the next few days and weeks. Otherwise, the outbreak in India can reach the level of the USA or Italy or could be worse than these countries within a few days or weeks, given the size of the population and lack of resources.

## Introduction

Ever since the outbreak of COVID-19, which was initially announced from China’s Wuhan city in December 2019 (1). The SARS-CoV-2 virus has affected more than 1,016,401 people in 202 countries and territories accounts for a total of 53,160 deaths worldwide, according to the Center for Systems Science and Engineering (CSSE) at Johns Hopkins University (JHU) and World Health Organization (WHO) (2,3). Therefore, a harmonized global response is desperately needed to prepare health care systems to coincide with this unprecedented challenge. Unfortunately, several countries like China, Italy, Iran, USA, Germany, Spain have been exposed to this disease drastically and passing critical lessons to contain the virus in the early stages to other countries. On April 2, India reported 2,586 confirmed COVID-19 cases followed by 73 deaths according to the http://gis.ndma.gov.in/arcgis/apps/sites/#/data and https://www.covid19india.org/ (4, 5). The number of confirmed cases seems less compared to other countries. However, taking into account the 1.3 billion population of India, it was necessary to take strict and essential measures as soon as possible. Within days of the epidemic, the Indian government ordered schools closure, religious events with large gatherings were either canceled or postponed. The local authorities encouraged residents to minimize the travel, avoid crowded places, and promoted physical distancing, travel restrictions imposed between states and cities, undoubtedly played a significant role in reducing the exposure and delayed the onset of the outbreak in several towns and villages in India.

In case, if COVID-19 spreads with the speed of Italy, Spain, China or the USA, it will become impossible to contain the disease without the discovery of vaccines, which will take at least 18 months from March 2020 according to the WHO report (6). The US has surpassed the position and recorded maximum cases of 216,721, including 5,137 deaths. Till 2 April 2020, Italy has endorsed 110,574 COVID-19 cases started with three cases in the first half of February, followed by 13,155 deaths to date according to https://coronavirus.jhu.edu/map.html (2). The primary reasons for the maximum death toll in Italy or other European countries like Spain are because of the slow response to the outbreak, delayed decisions on travel ban, and locking down the country. Italy is the second-largest country with the oldest population in the world, lack of testing kits, lack of social distancing practice due to the family cultures. However, China’s Wuhan region was locked down since mid-January from the rest of the world, and they have not reported any new positive cases for many days. Interestingly, it seems like strict lockdowns appear to have an extraordinary impact on controlling the disease (7).

At the instant, our national health system’s potential to adequately react to the requirements of those who are already infected and demand serious attention. India needs to speed up the diagnosis procedure because until 31 March 2020, by officials Indian Council of Medical Research (ICMR), only 42,788 individuals were tested for COVID-19, which is relatively a minimal number compared to the total population of India. The capacity to perform tests on 1.3 billion people is still a question. Alternatively, the only way to minimize unnecessary deaths is to extend the period of country lockdown for the next few weeks. Studies reported on 1918–19 pandemic suggest that holds and delays in proposing social distancing actions are associated with excess fatality (8, 9). Another major issue in India is more than half of the population lives on daily wages who require the basic need of living that includes food and shelter. Extending the lockdown period will have a significant impact on such vulnerable populations in the country. We suggest the Indian government consider fulfilling basic requirements that include food and shelter. Our analysis verifies the necessity and demand for continued reinforcement of mitigatory social and group distancing.

### Forecasting

We provide the platform to our political leaders and the government for preparation through forecasting the situation that could happen in upcoming days. It will help them to make adequate measures plus create the appropriate facility accordingly for the COVID-19 infected people. According to our study and the available data, while China’s new cases follow an almost flat curve, other countries such as India, US, Italy, Spain, and Germany still have exponential growth. Particularly to point India’s exponential growth followed by Spain and Germany is much worse than Italy besides the number of total cases because of the steep slope of growth rate (Figure 1).

**Figure 1.**
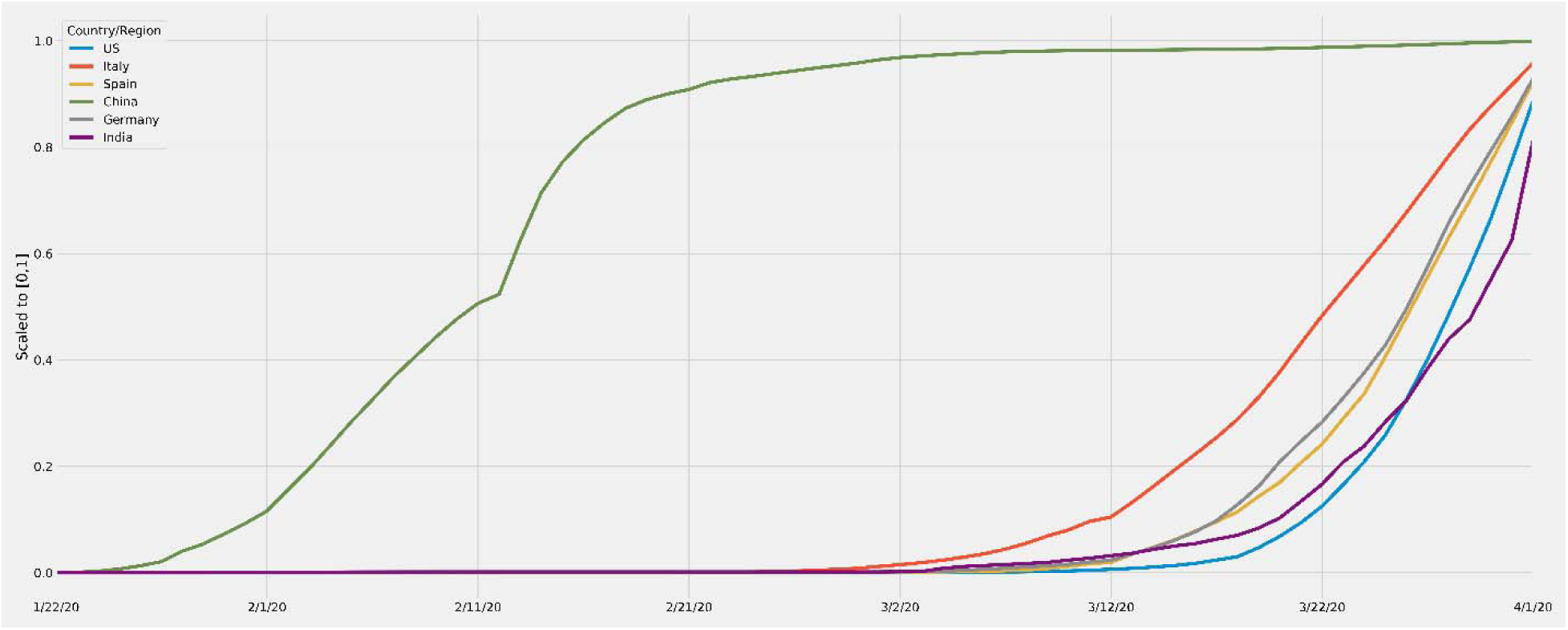
COVID-19 growth rate: Growth rate comparison of five countries (US, Italy, Spain, China, Germany) with India to determine rate of infection in the near future.

On the other hand, the US is inferior. We kept the frequency of all countries scaled to [0, 1] range only for the trend comparison. As the Indian COVID-19 cases are in exponential growth rate, it is expected that a magnitude of the number of infected cases is coming in the future.

On 1 April, the total number of tested individuals was 47951, in which 1637 are positive cases, which is about 3.5% of the total tested individuals according to the crowdsourced patient database (5). This rate is increasing with new test results coming day by day. Figure 2 depicts the moving average percentage, with a span of 7 days, of total positive cases.

**Figure 2.**
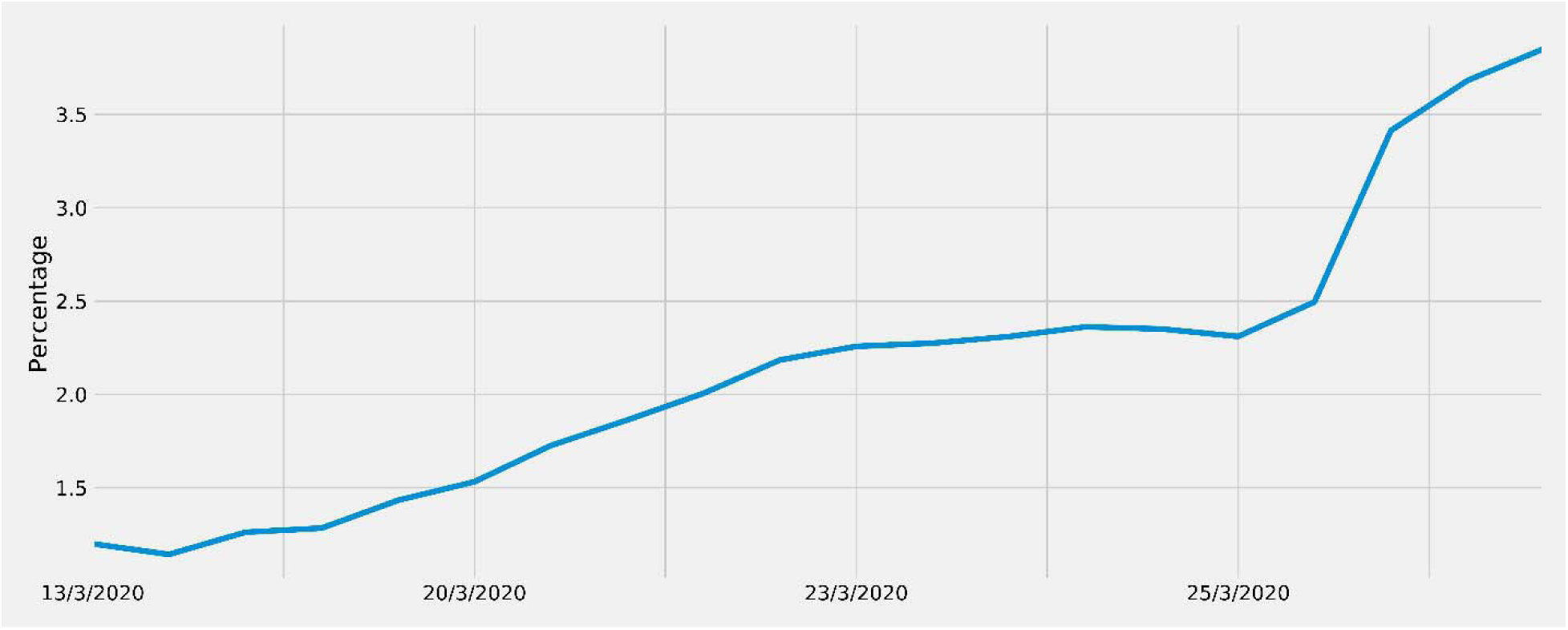
Moving average curve for tested samples: Percentage moving average of confirmed cases out of the total tested individuals per day. The slope describes the moving average percentage, with a span of 7 days, of total positive cases.

The central government of India took the lockdown decision is the crucial step; however, it is hard to say that the lockdown of 21 days is enough. As mentioned above, SARS-CoV-2 infection takes around 2-14 days to show its symptoms, and the cases are increasing with new tests on individuals. It raises the concern that how many days these numbers will go up and what quantity of arrangement is required for upcoming days.

Based on the given crowdsourced patient database from 31 January, it is possible to estimate the outcomes of future days and weeks. To predict when the exponential growth will become linear and then flat, we use the growth factor of new cases per day. The 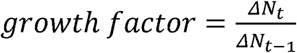; where *N*_*t*_ is the number of total positive cases on t day and *N*_*t*−1_ is the number of total cases on the previous day. In simple terms, it is the ratio of new cases on t day and new cases of the previous day. Figure 3 (left) depicts the moving average growth factor of recent days with a span of 7 days. Figure 3 (Right) illustrates the predicted growth factor with a standard deviation of 0.3443 for the upcoming days. When the growth factor becomes 1.0, we can say that the rate is no longer exponential. It is also known as the Inflection point. The estimated time to reach the inflection point is between 15-20 April. At that time, the estimated number of total positive cases will be over 12500, if lockdown remains continue.

**Figure 3.**
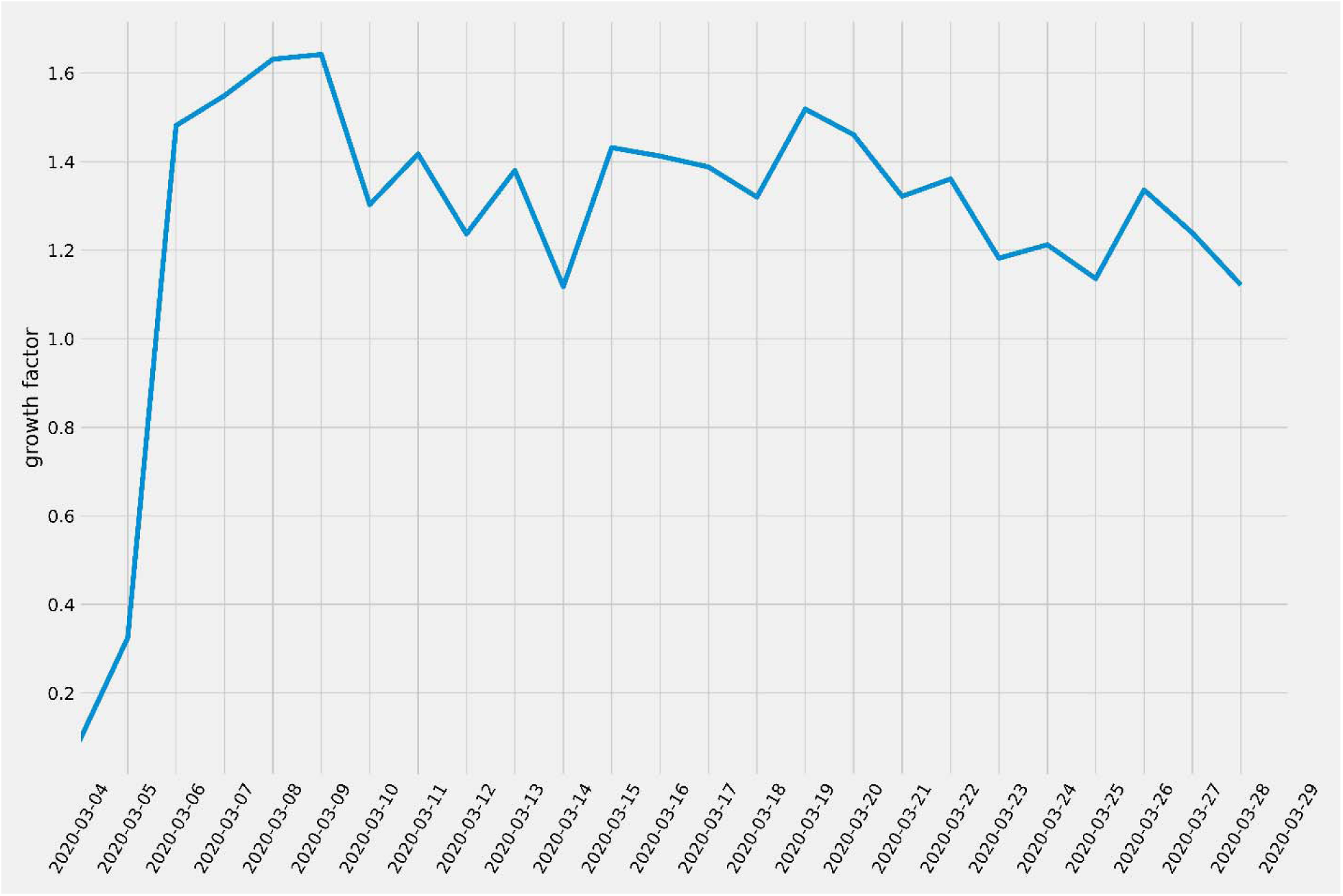
Growth factor prediction for present and future: The moving average of the growth factor determined A) Based on the available previous days data

**Figure 3.**
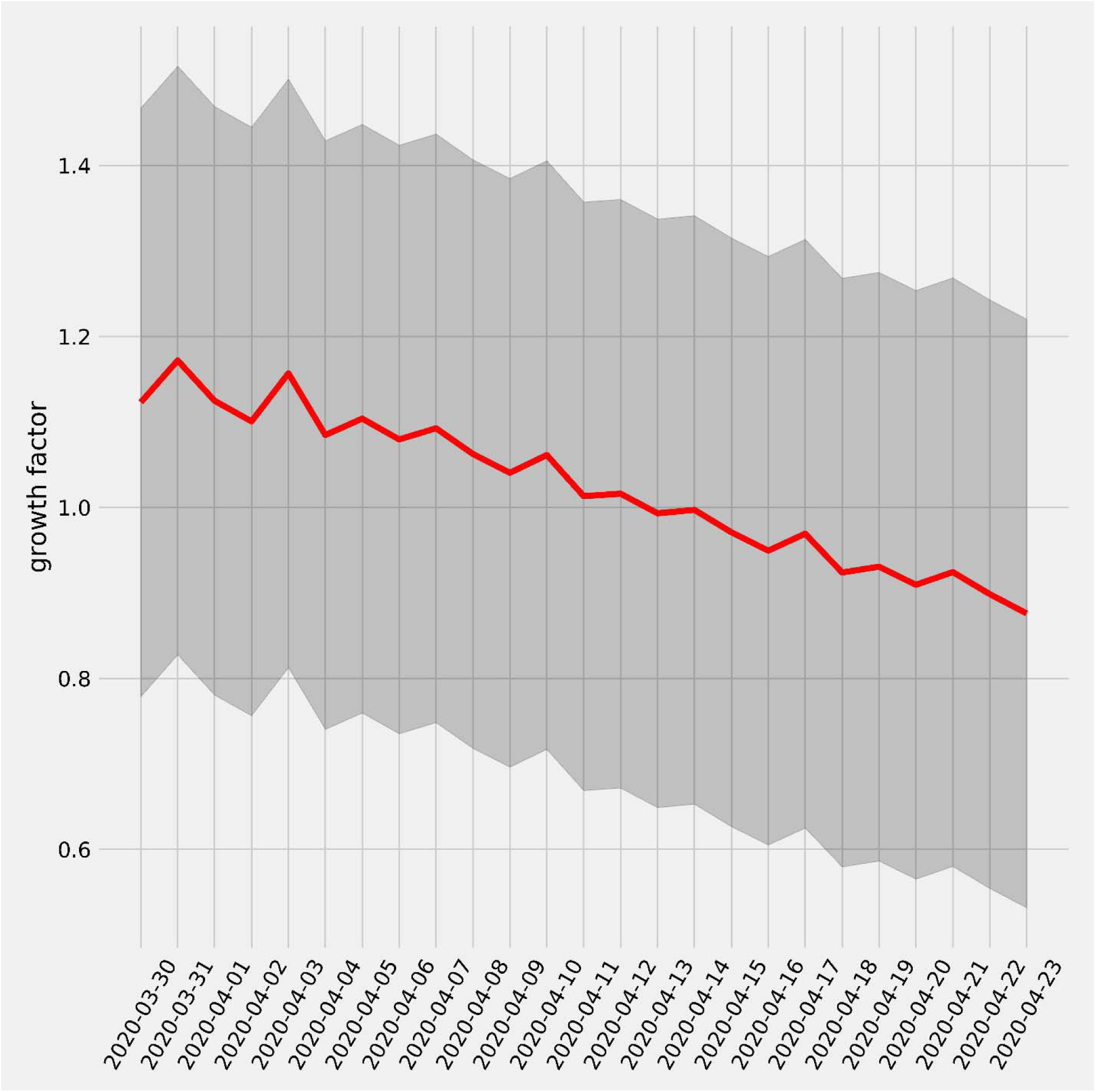
Growth factor prediction for present and future: The moving average of the growth factor determined B) The upcoming days forecast till 15 April.

Once the inflection point is obtained, the further curve will follow the logistic curve, as shown in Figure 4. After this point, the number of infected cases will still increase, but the rate will no longer be exponential. Eventually, it will become flat then decrease. But, these results correspond to the lockdown. As suggested by Singh and Adhikari (10), the lockdown must extend further to 13-15 May. As a result, our predictions are in support of the same. After 49 days, the estimated number of positive cases will be over 20000. If the lockdown continues till 13-15 May, the estimated number of new cases will be less than 100.

**Figure 4.**
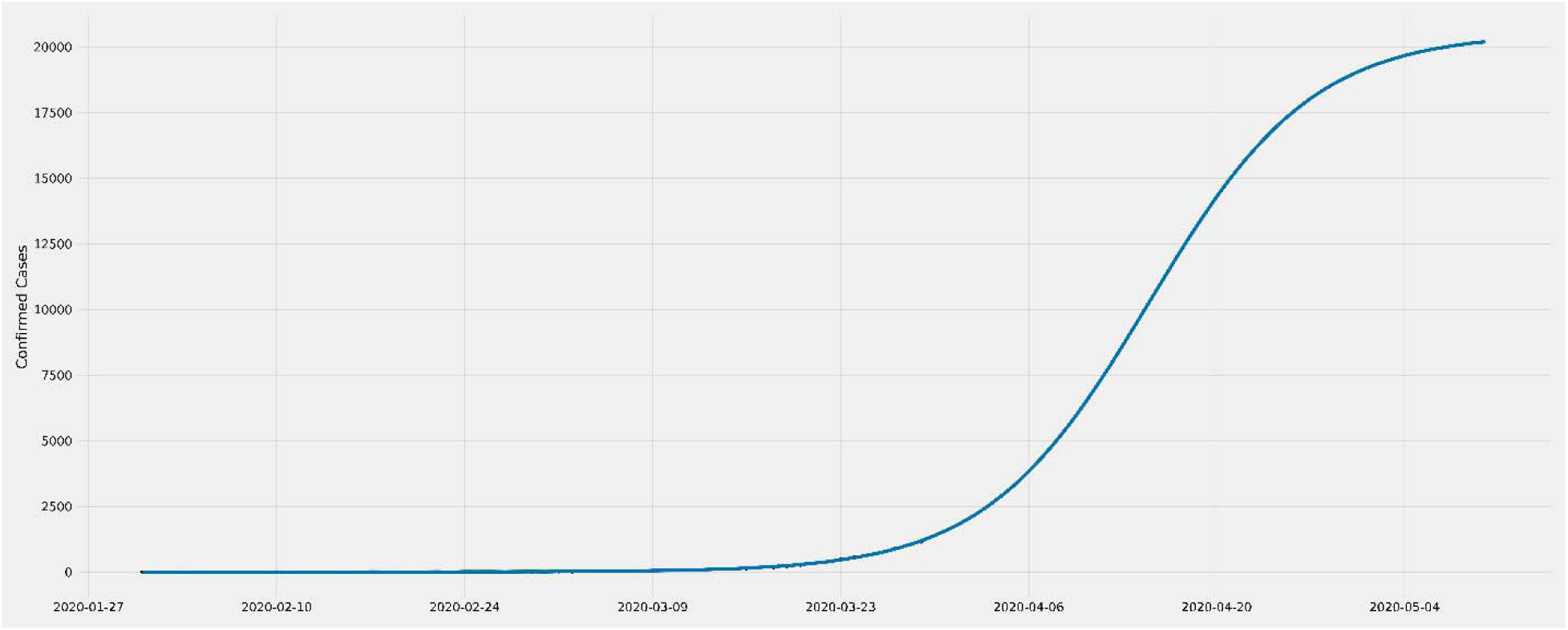
Logistic curve: This determine the identification of the inflection point and when the rate will no longer be exponential. It also shows predictions of each day. The dotted line represents the time point of the infection outbreak in Italy. It is expected that the number of cumulative patients who are infected will start to deviate from the exponential low in 3–4 days. The plateau of the cumulative curve will be reached just over 30 days from March 11, 2020.

We use Lasso Regression with *α* = 0.12 and Polynomial features of degree 2 to predict the growth factor, Figure 3 (Right), by fitting the moving average values of given data of recent days. The Logistic curve in Figure 4 is predicted using the Prophet Python module provided by Facebook. The growth is set to logistic, and carrying capacity is 20000, which is the estimated maximum confirmed cases till 15 May. These maximum cases are calculated using the predicted growth rate and the formula given later in this section to compute new cases per day.

Given all the information above, it is also possible to estimate the number of cases on the next day. This estimate is valuable to prepare our National Health systems and government for the expected catastrophe. These numbers of expected confirmed cases can aid the Indian intensive care facilities and hospitals to be ready ahead of time. The formula to estimate the minimum and maximum number of new cases on a day is

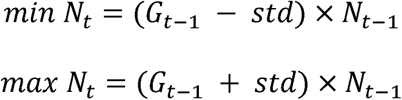

*G* is the growth factor, std is 0.3443 (computed earlier), and *N*_*t*−1_ is the number of cases on t-1 day.

## Discussion

Extraordinary measures required to handle challenging circumstances. Severely affected countries have passed an important message to take early strict actions than dealing with consequences once the SARS-CoV-2 has spread. On 24 March 2020, the Indian government has taken decisive action to interrupt the chain of COVID-19 transmission. The order was to shut down the schools, universities, workplaces and put the “Janata curfew” to lock down the whole nation in the attempt to complete the cessation of public contact for brief and extended periods of 21 days.

India being a republic, secular, multi-religious country, it is difficult to impose the lockdown overnight without proper awareness and announcements (11). Therefore, Government and local authorities need to take further strict actions and appropriately observe religious events and large gatherings beforehand to stop the uncontrollable spread of COVID-19. Further, it is essential to focus on measures like self-isolation, social distancing, testing all of the suspected cases, and increase the hospital facilities to treat COVID-19 patients as much as possible.

It is found that SARS-CoV-2 infection has a more severe impact on older people and individuals with medical history or ongoing conditions such as diabetes, high blood pressure, Cardiovascular disease (CVDs), cancer, or other serious illnesses (12, 13, 14). In Italy, where more than two-thirds of patients died with the mean age of 81 years, reported to be former smokers and suffering from various comorbidities. It clears the fact that having these situations causes acute respiratory distress syndrome (ARDS) by SARS-CoV-2 pneumonia, requires a respiratory support system, and would not have died otherwise. The estimates suggest that more than 72 million people have diabetes, 54.5 million CVDs cases in the adult population of India (15, 16). In this position, the most challenging enigma is to divine the cumulative number of infected patients. The first approach should be to identify the infected individuals in India; however, because Indian hospitals are not able to test the sufficient number of patients so the actual cases might be much higher than the reported number. According to our analysis of the moving average curve to date 27 March 2020 in figure 2, it is inevitable that India has many more cases to be confirmed as the hospitals will perform more tests in the coming days. Beyond this, it is critical to find how many of them will need intensive care following admission.

Our prediction is of crucial importance to prepare for extra facilities in Indian hospitals and to measure the period in which they need to be ready. If the lockdown is not going to extend, it might not be unrealistic to assume what is going to happen in India within days or weeks, might be a reflection of what happened in the USA and Italy. Unfortunately, we don’t have additional evidence that can take into consideration to identify the exact number of infection cases, recovery, and deaths in India. Our prediction is till 30 April 2020 and is based on the basis available data recorded until 02 April 2020. It is important to note that this prediction is based on the lockdown data, which is applicable until 14 April 2020. If the lockdown will not extend according to the predicted number of days by (10), without further data, it is difficult to predict the situation.

The only possibility for India is to learn from China’s strict precautionary measures where the more affected regions were taken over by the military and quarantining everyone, even with minor symptoms and people who came in contact with an infected individual. By applying these measures, the Hubei region has recorded zero number of newly infected cases after about two months of strict shutdown. Keeping in mind the population of Hubei, which is 50 million in comparison to the 1,030 million Indian population, it requires a longer lockdown than 21 days. However, another challenge for the Indian Government is to tackle millions of destitute people who lack food and proper shelter to perform self-isolation practices.

There is an urgent need to test maximum numbers of people, especially in the whole infected region of the countries such as New Delhi, Maharashtra, and Tamil Nadu, to accumulate the knowledge warranted at this time. It is noteworthy to describe that without additional data, it is unmanageable to execute further robust deductions regarding the thorough estimate of patients who will get infected soon. The intention immediately is to raise this figure to satisfy pressing coming demands securely. We have a few weeks to accomplish this goal in terms of obtaining personnel, technical equipment, and materials. These problems are majorly applicable to other underdeveloped or developing countries where a suitable quantity of intensive care facilities are not available and not feasible to produce in such a short period. In this case, the condition might spread out of control and improbable. According to our forecast, an inflection point is between 15-20 April. It defines that cases in India will remain constant, and no further addition of new cases will be reported from this point, but keeping this in mind that this point is only possible if lockdown will continue and testing all the possible suspects.

## Conclusion

The COVID-19 outbreak in India is relatively new compared to other countries, proper surveillance, transparent reporting of characteristics of infected individuals is necessary to update regularly. Nevertheless, aggressive steps need to be taken with critically ill SARS-CoV-2 infected patients, including isolation and ventilatory support. Our findings represent further lockdown restrictions that need to be extended urgently, in a matter of days. To tackle the substantial number of unnecessary deaths, we need to focus on increasing the testing capacity and speed up the rate of recovery of infected people by providing all the necessary support and medication. For individuals with other comorbidities or older people requires regular testing, more robust personal protection and should have access to intensive surveillance or therapy.

## Data Availability

All data is found within the manuscript or publicly available through the cited web sources

## Authors’ contributions

GK and SAA wrote the manuscript and designed the concept, JSV performed the prediction analysis. JSK, SAA, and GK contributed to the final correction of the manuscript. All authors read and approved the final manuscript.

## Acknowledgments

GK is thankful to University of New South Wales, UNSW Sydney Australia for the Research fellowship. SAA is thankful to the Indian Council of Medical Research (ICMR), India for the Senior Research fellowship. JKV is thankful to Indian Institute of Science for the Research fellowship.

## References

1. Zhu N, Zhang D, Wang W, Li X, Yang B, Song J, Zhao X, Huang B, Shi W, Lu R, Niu P. A novel coronavirus from patients with pneumonia in China, 2019. New England Journal of Medicine. 2020 Jan 24.

2. Dong E, Du H, Gardner L. An interactive web-based dashboard to track COVID-19 in real-time. The Lancet Infectious Diseases. 2020 Feb 19 Johns Hopkins Center for Systems Science and Engineering. Coronavirus COVID-19 Global Cases. 2020. https://systems.jhu.edu/research/public-health/ncov/ (accessed 2 April 2020).

3. World Health Organization (WHO) https://experience.arcgis.com/experience/685d0ace521648f8a5beeeee1b9125cd (accessed 2 April 2020)

4. http://gis.ndma.gov.in/arcgis/apps/sites/#/data (accessed 2 April 2020)

5. https://www.covid19india.org/ (accessed; 2 April 2020)

6. Landscape of COVID-19 candidate vaccines (PDF). WHO. 20 March 2020 https://www.who.int/blueprint/priority-diseases/key-action/novel-coronavirus-landscape-ncov.pdf?ua=1

7. Fong MW, Gao H, Wong JY, Xiao J, Shiu EY, Ryu S, Cowling BJ. Nonpharmaceutical Measures for Pandemic Influenza in Nonhealthcare Settings-Social Distancing Measures. Emerging infectious diseases. 2020 May 17;26(5).

8. Bootsma MC, Ferguson NM. The effect of public health measures on the 1918 influenza pandemic in US cities. Proceedings of the National Academy of Sciences. 2007 May 1;104(18):7588–93.

9. Hatchett RJ, Mecher CE, Lipsitch M. Public health interventions and epidemic intensity during the 1918 influenza pandemic. Proceedings of the National Academy of Sciences. 2007 May 1;104(18):7582–7.

10. Singh R, Adhikari R. Age-structured impact of social distancing on the COVID-19 epidemic in India. arXiv preprint 2003.12055. 2020 Mar 26

11. Coronavirus in India: Tablighi Jamaat preacher, others booked for violating govt guidelines on religious gatherings (https://www.indiatoday.in/india/story/coronavirus-in-india-tablighi-jamaat-preacher-others-booked-for-violating-govt-guidelines-on-religious-gatherings-1661870-2020-03-31) (accessed 2 April 2020)

12. Hill MA, Mantzoros C, Sowers JR. Commentary: COVID-19 in Patients with Diabetes. Metabolism-Clinical and Experimental. 2020 Mar 24.

13. Tan W, Aboulhosn J. The cardiovascular burden of coronavirus disease 2019 (COVID-19) with a focus on congenital heart disease. International Journal of Cardiology. 2020 Mar 28.

14. Lake MA. What we know so far: COVID-19 current clinical knowledge and research. Clinical Medicine. 2020 Mar;20(2):124.

15. Diabetes is India’s fastest growing disease: 72 million cases recorded in 2017, figure expected to nearly double by 2025. https://www.firstpost.com/india/diabetes-is-indias-fastest-growing-disease-72-million-cases-recorded-in-2017-figure-expected-to-nearly-double-by-2025-4435203.html. (accessed 2 April 2020)

16. Abdul-Aziz AA, Desikan P, Prabhakaran D, Schroeder LF. Tackling the burden of cardiovascular diseases in India: The essential diagnostics list. Circulation: Cardiovascular Quality and Outcomes. 2019 Apr;12(4):e005195.

